# Validation of virtual trichrome stains for kidney fibrosis evaluation using dual-mode emission and transmission microscopy

**DOI:** 10.1101/2025.11.11.25339911

**Authors:** Willy Ju, Samuel Border, Fatemeh Afsari, Srishti Seth, Sarah Rezapourdamanab, Ruben Renteria, Shalini S. Ramachandra, Rajib Gupta, Fadi Salem, Alton B. Farris, Richard Levenson, Jarcy Zee, Pinaki Sarder, Kuang-Yu Jen, Farzad Fereidouni

## Abstract

Assessment of interstitial fibrosis is essential in the diagnosis and prognosis of kidney diseases. However, current histologic scoring methods using trichrome-stained slides are limited by inter-observer variability and inconsistent stain reproducibility. To address these challenges, we developed DUET (DUal-mode Emission and Transmission) microscopy, a novel imaging platform that rapidly captures both brightfield and fluorescence images from H&E-stained slides to generate pixel-registered collagen images and virtual trichrome stains. In a cohort of 32 kidney transplant biopsies, four renal pathologists estimated the extent of interstitial fibrosis in real trichrome and DUET-derived virtual trichrome whole slide images, with the latter showing improved inter-pathologist agreement. A deep learning pipeline was trained to segment interstitial kidney regions from DUET-acquired images, enabling semi-automated computational fibrosis quantitation, which demonstrated a positive correlation with pathologists’ estimates of interstitial fibrosis. These findings highlight DUET as a rapid, cost-effective, and scalable alternative to traditional trichrome staining, offering both visual and computational advantages.

## INTRODUCTION

Interstitial fibrosis (IF) is a common process of chronic kidney injury in all renal diseases [1]. It is a key feature that renal pathologists routinely report to convey an estimated extent of irrecoverable renal injury, which has prognostic value for predicting the trajectory of future renal function. IF involves an excess accumulation of extracellular matrix that is primarily composed of collagen and, in essence, represents the extent of scar tissue within the kidneys. A large body of literature supports the strong association between the degree of IF and renal outcomes [2-5]. As such, estimation of IF has been formally incorporated into guidelines for the classification of several renal diseases, including the Oxford Classification for IgA nephropathy [6], the Banff Classification for transplant rejection [7], and the International Society of Nephrology/Renal Pathology Society Classification for lupus nephritis [8, 9].

Currently, it is standard practice for pathologists to use trichrome-stained slides to visually estimate the extent of IF. The trichrome stain helps to highlight collagen and is typically preferred over other histochemical stains. However, studies have shown that pathologists’ microscopic evaluation of IF is often hampered by problems with reproducibility [10]. The trichrome stain is neither specific nor sensitive for collagen, leading to both underestimation of collagen content in cases of subtle or early fibrosis and overestimation of collagen content by nonspecifically staining interstitial edema or other structures [10-13]. Technical variability of trichrome stains between laboratories as well as within the same laboratory can also affect the reproducibility of fibrosis estimation [14].

Recent efforts to create virtual trichrome stains using generative adversarial networks (GANs) have demonstrated the potential to reproduce trichrome-like appearance from standard H&E slides, with pathologist validation [15, 16]. However, these methods typically require large, high-quality training datasets and can be susceptible to artifacts or hallucinated structures inherent to generative modeling.

To improve collagen visualization, we have developed a novel microscopy approach termed DUET (Dual-mode Emission and Transmission). This whole-slide imaging technique rapidly scans H&E-stained slides in both brightfield and fluorescence modalities, extracting the signal for collagen from the fluorescence image within minutes. These high-fidelity collagen signals can then be incorporated into the H&E images to generate virtual trichrome images, eliminating the need for additional stains or labeling steps [17].

In this study, we apply DUET to transplant kidney biopsies, producing a collagen map and virtual trichrome whole-slide images that renal pathologists assessed to demonstrate the practical utility of this imaging technique. Furthermore, we developed a semi-automated computational pipeline for estimating IF in these kidney biopsies using the collagen image extracted from DUET data. Our approach aims to address the challenges of traditional clinical and computational fibrosis estimation by deploying a whole slide-capable, accurate, quantitative, and reproducible method for assessing IF in kidneys, leveraging complementary information from both brightfield and fluorescence imaging modalities.

## METHODS

### DUET Image Acquisition

Pixel-registered brightfield and fluorescence whole-slide images were acquired from formalin-fixed paraffin-embedded (FFPE) hematoxylin and eosin (H&E) stained slides using the Dual-mode Emission and Transmission (DUET) imaging system. For fluorescence imaging, epifluorescent excitation was employed with a 405 nm LED light source (LED Engin, LZ1-00UB00), directed toward the specimen through a broadband dichroic mirror (Semrock, Di03-R405-t1-25 × 36) and focused using a 20× objective lens (Nikon, NA = 0.75). The emitted fluorescence signal was captured with a 9-megapixel scientific-grade CCD color camera (Ximea, MD091CU-SY) coupled to a 200 mm tube lens (Thorlabs, ILT 200). Both brightfield and fluorescence channels were acquired using an automated XYZ motorized stage (Zaber, X-ASR050B050B-SD12 and X-DMQ12L-AE55D12), controlled by custom software written in VB.NET.

### Case Selection

32 core needle transplant kidney biopsy cases with a range of IF were selected from the clinical archive. Of the 32 cases, 23 were sourced from the original H&E and trichrome slides used for routine clinical care, sectioned and stained by the UC Davis Health Pathology Laboratory. The remaining 9 cases were generated from the original tissue blocks but stained in a different research laboratory (UC Davis Center for Genomic Pathology Lab). Ethics Statement This study was conducted in accordance with the principles of the Declaration of Helsinki and was approved by the Institutional Review Board (IRB) at the University of California, Davis (Protocol #2013364). The use of archived, de-identified human tissue specimens was determined to qualify as human subjects research with a waiver of informed consent, as the study posed minimal risk and involved no direct patient contact.

### Automated Collagen Mask Extraction Using Deep Learning

The fluorescence image contains clear signals corresponding to basement membranes and bulk collagen surrounding glomeruli renal tubules, features composed mainly of collagen types III and IV(Figure 1). These basement membrane structures are critical for evaluating tubular atrophy and interstitial expansion, both of which are central to fibrosis scoring. However, such features are often difficult to resolve using standard brightfield imaging alone. By capturing these signals in the fluorescence channel, DUET enables more precise localization of collagen within the tissue architecture, especially in areas where fibrosis may be subtle or early.

**Figure 1.**
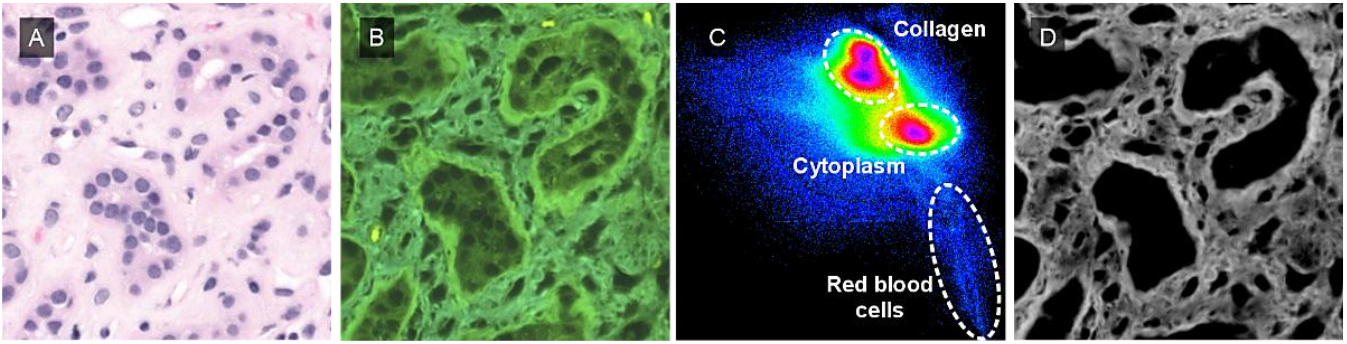
Generation of collagen ground-truth labels using a spectral phasor approach. (A) Representative DUET fluorescence image of an H&E-stained kidney section. (B) Spectral phasor plot showing distinct clusters corresponding to collagen, cytoplasm, and red blood cells used to identify collagen-positive pixels in the fluorescence image. (C) A collagen mask derived from the spectral phasor analysis was subsequently used as ground truth for training the automated collagen extraction. Scale bar: 50 μm (applies to all images).

In order to automate the process, ground truth labels were generated using our previously published spectral phasor approach [17-19], as shown in Figure 1. The spectral phasor approach allows to highlight collagen and cytoplasm based on the color difference between collagen and cytoplasm on fluorescence images. The segmentation was performed automatically by identifying the individual clusters and performing a reciprocal transformation to find the pixels associated with each cluster.

The outcome of the segmentation was supervised and necessary minor manual adjustment was performed to ensure the maximum segregation. One of the slides was imaged using second-harmonic generation (SHG) to verify the collagen ground-truth labels.

700 image patches from three cases with a different range of fibrosis were used and the output of clustering and brightfield and fluorescence images were used for training Unet++ model. The input dimensions for training were 512×512 pixels. A UNet++ model architecture was employed, utilizing the Adam optimizer with an initial learning rate of 5e-5 and 1000 training steps, with a batch size of 2. The training images comprised 80% of the original annotated image dataset.

### Virtual Trichrome Creation

We used our deep learning model to segment collagen mask from the fluorescence channel. Next, the collagen mask was pseudo-colored and overlaid onto a synthetic trichrome base derived from the brightfield image. Figure 2 illustrates this process, showing the brightfield and fluorescence inputs, the intermediate collagen mask and base image, the final virtual trichrome, and a real trichrome image from a serial section for comparison. Unlike conventional trichrome stains, the virtual trichrome maintains nuclear detail and microanatomic clarity, and can be generated in whole-slide format for direct comparison with the original H&E layer.

**Figure 2.**
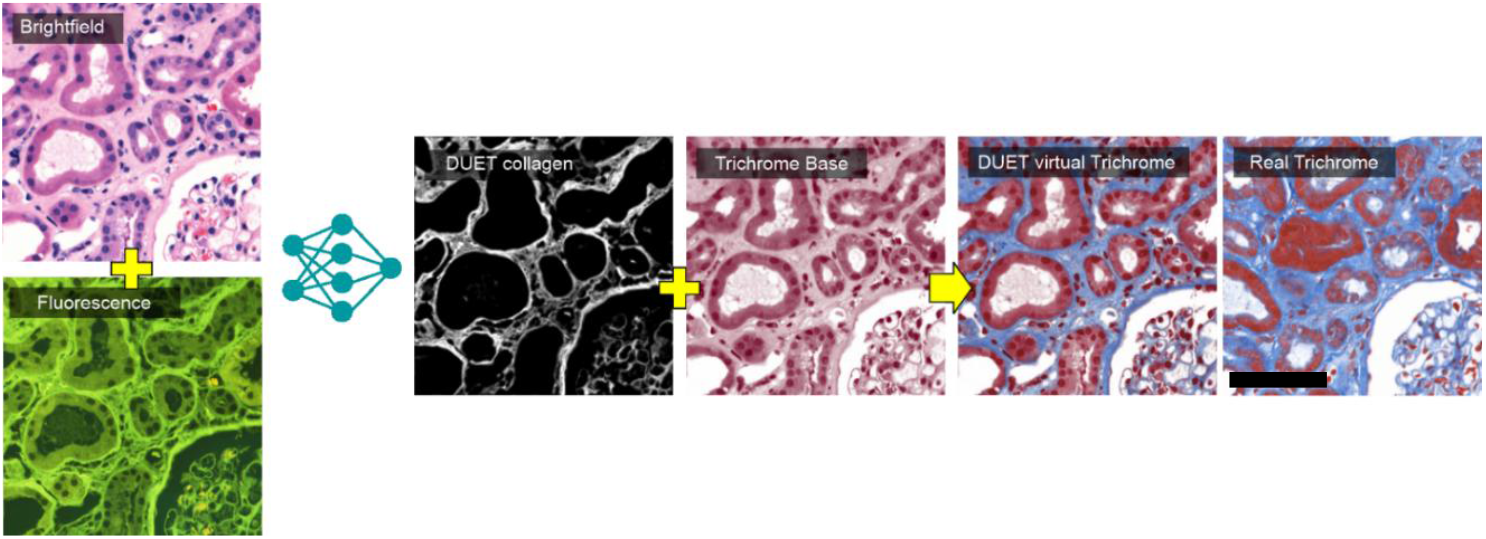
Virtual trichrome images were generated using pixel-registered brightfield and fluorescence images acquired from the same H&E-stained slide as inputs. A collagen mask was extracted from the fluorescence image using DUET and a deep learning model. A trichrome base image was synthesized from the brightfield channel, and the DUET-derived collagen mask was overlaid onto this base to create the virtual trichrome. For comparison, a real trichrome image from a serial section of the same tissue region was acquired using a conventional brightfield scan. Scale bar: 100 μm (applies to all images).

### Computational Fibrosis Quantitation

Computational IF quantitation was calculated using DUET-extracted collagen-positive pixels. Glomeruli, arteries, and non-cortical regions were excluded from the analysis using pathologist-guided annotations. The computational IF quantitation was calculated as the proportion of DUET-extracted collagen-positive pixels relative to the total cortex area. Computational fibrosis quantitations were then correlated to the pathologists’ visual estimates.

### Pathologist Fibrosis Estimation

Four renal pathologists estimated the percentage of renal cortical IF involvement to the closest 5% on 32 real trichrome whole slide images (set 1) and their corresponding virtual trichrome whole slide images (set 2). The order of the cases for each set were randomized, and a 2-week washout period was implemented between evaluation of the sets. The virtual trichrome images were generated from the H&E slides in closest proximity to the real trichrome slides in terms of depth within the paraffin block.

### Statistical Analysis

Since each sample was rated twice per pathologist, once for real trichrome and once for virtual trichrome, the intraclass correlation coefficient (ICC) was calculated within subgroups. The ICC was calculated as a ratio of between-sample variability to the sum of between- and within-sample variability, based on a random effects model, and is therefore bounded by zero. 95% parametrically bootstrapped confidence intervals were calculated and reported for each ICC using the bootMer function in the lme4 R package with 1000 iterations. Additional analysis was conducted to compare the coefficients of determination (R^2^) for each pathologist’s IF estimates with the average estimates across all pathologists, for both real and virtual trichrome images.

## RESULTS

### Performance of Automated Collagen Signal Processing from DUET Images

Collagen prediction images were generated as 2D masks, which were then binarized before calculating segmentation metrics, which include the AUC, Sensitivity, Specificity, Dice score, and Accuracy (shown in Table 1). Examples of predicted collagen images from various kidney features, including tubules, interstitium, glomerulus, inflammation, and artery, are compared to the corresponding brightfield and fluorescence images in Figure 3.

**Table 1.**
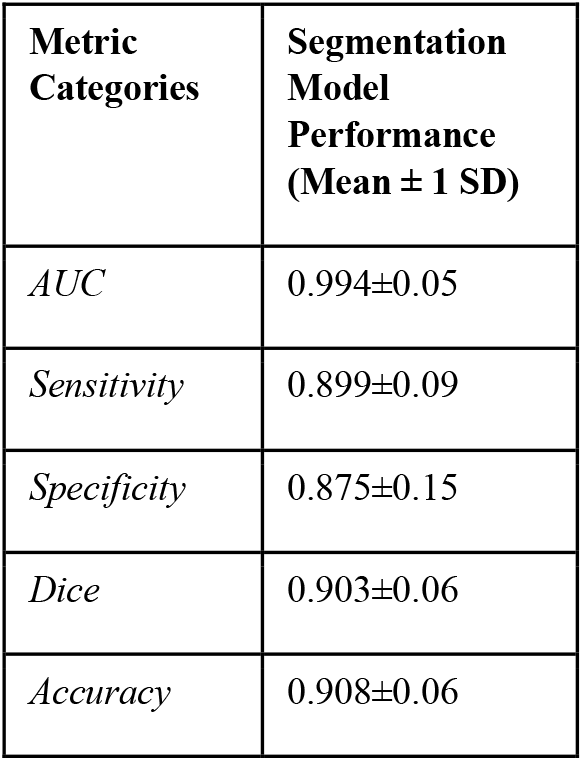
Performance metrics for collagen segmentation using concatenated brightfield and fluorescence inputs are shown (AUC, Sensitivity, Specificity, Dice, Accuracy).

**Figure 3.**
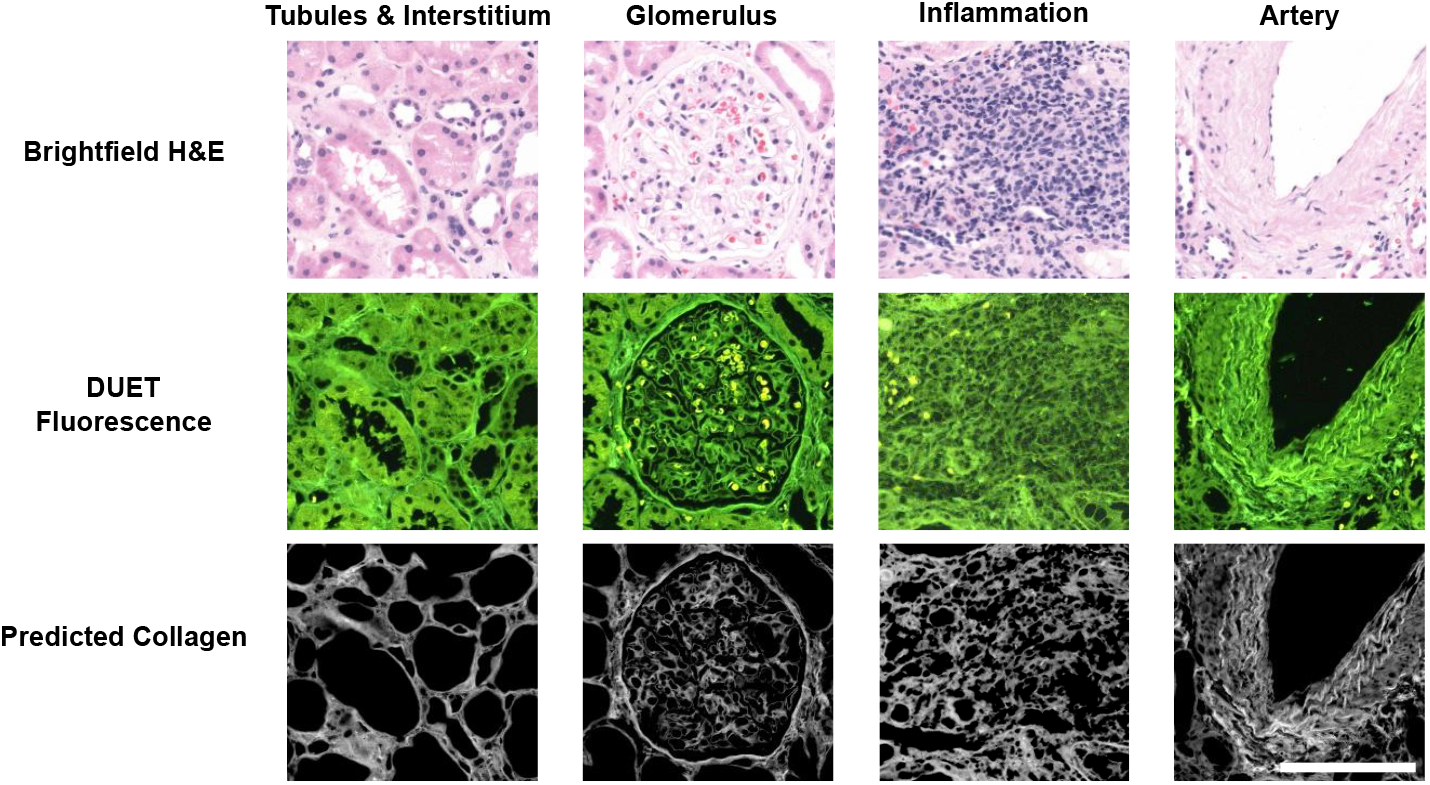
Example 512×512 image patches of brightfield and fluorescence inputs shown with predicted collagen masks of different kidney features, including: Tubules & Interstitium, Glomerulus, Inflammation, and Artery. Scale bar: 100 μm (applies to all images)

### Virtual Trichrome Evaluation

From the DUET-derived collagen signals, virtual trichrome images were created. We assessed the consistency of virtual trichrome images under variable staining conditions by comparing H&E and trichrome-stained sections from two different laboratories using serial sections from the same kidney specimen (Figure 4). Qualitatively, the color profiles of both the H&E (Figure 4A and 4E) and real trichrome stains (Figure 4B and 4F) are markedly different. Lab A’s trichrome shows stronger contrast between fuchsinophilic (red-staining) tubules and blue-staining collagen, whereas Lab B’s stain provides better spatial clarity and improved glomerular and tubular basement membrane staining. In contrast, the virtual trichrome images generated from DUET remain visually consistent across both sources. Despite variation in the original H&E stains, the virtual trichrome images preserve similar color contrast and maintain detail within the basement membrane (Figure 4C and 4G). Notably, the collagen distribution extracted from DUET fluorescence images retains reliable structural quality, unaffected by differences in the initial H&E stain appearance (Figure 4D and 4H).

**Figure 4.**
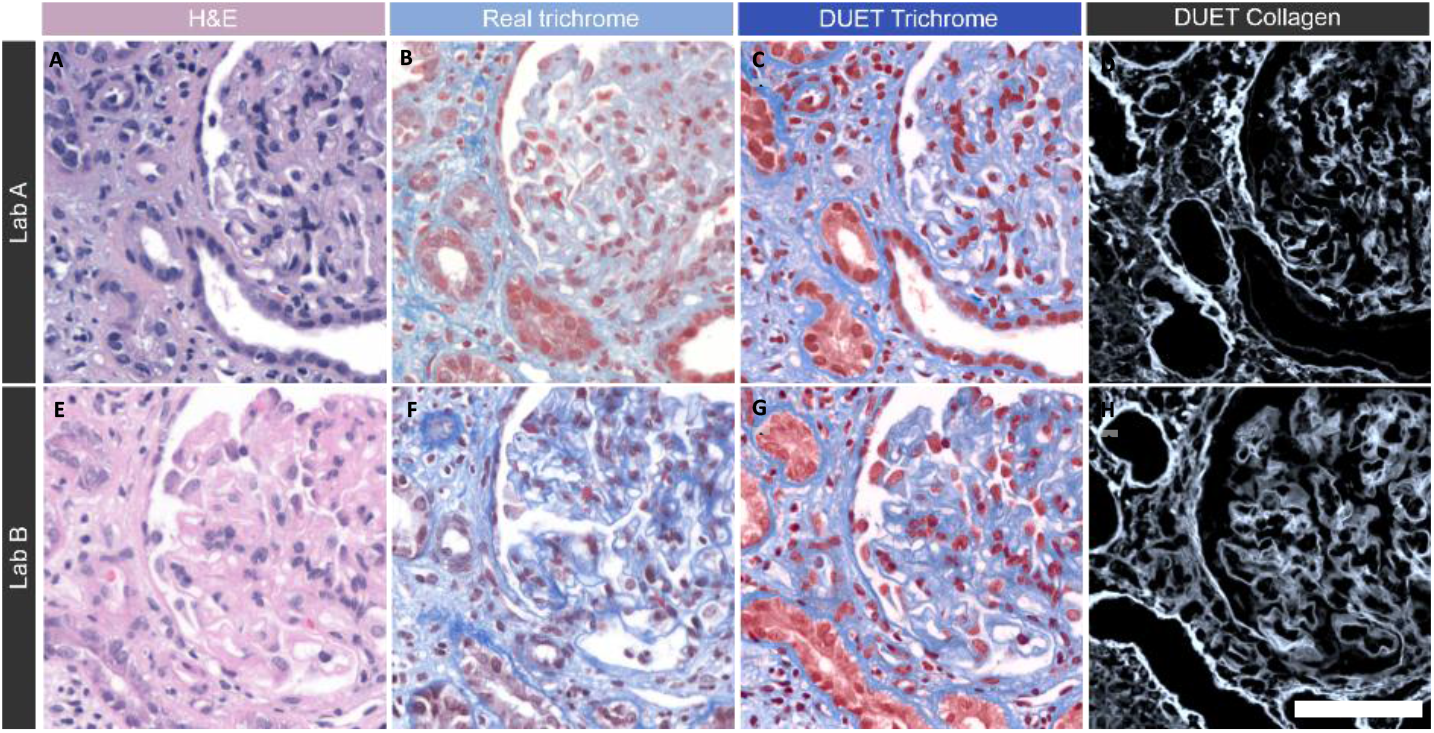
Representative kidney H&E (A,E), real trichrome (B,F), DUET trichrome (C,G), and DUET-extracted collagen (D,H) images from two different labs (Lab A, row 1; Lab B, row 2) at UC Davis. The Lab A trichrome (B) shows stronger contrast (red tubules vs turquoise collagen), whereas the Lab B trichrome (F) shows better spatial resolution and specific binding. For example, the basement membrane is visible around tubules, and capillary structures are more distinct inside the glomerulus (F). In contrast, the DUET trichrome (C,G) provides a consistent color theme and quality across both H&E sources. Collagen distributions extracted from the H&E images (D,H) remain of similar quality despite differences in staining. Scale bar: 100 μm (applies to all images).

Figure 5 compares real and DUET-derived trichrome stains. Traditional trichrome images at low (A) and high (C) magnification are compared to the corresponding DUET virtual trichrome images (B, D), demonstrating improved color contrast, structural detail, and visualization of collagen.

**Figure 5.**
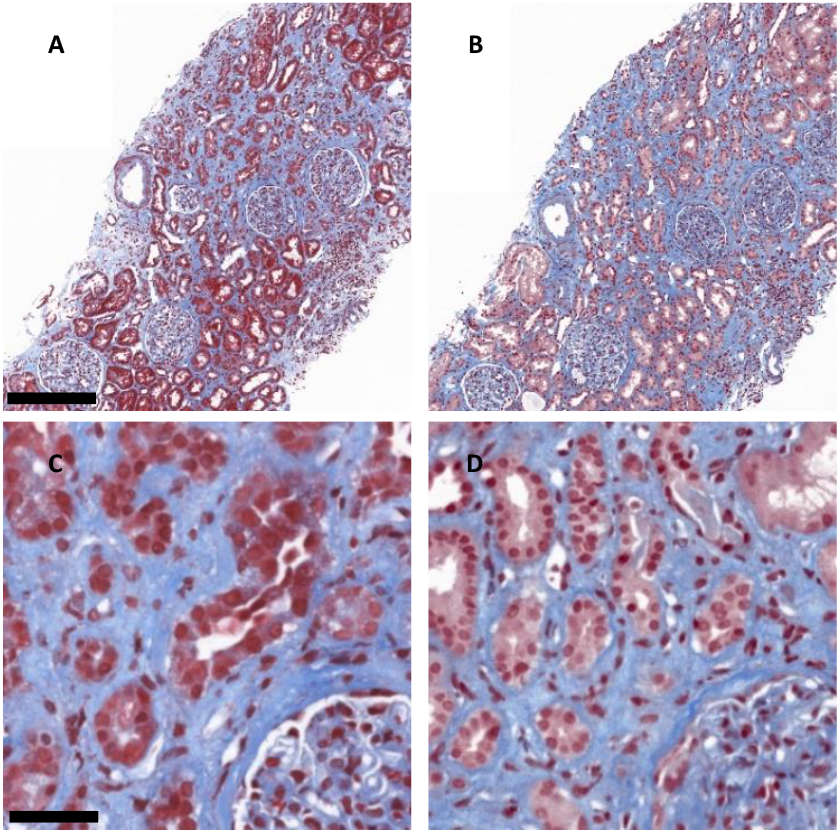
Low- and high-power comparisons of real and DUET-derived trichrome stains from the same kidney biopsy region. Panels A and C show traditional trichrome stains highlighting interstitial collagen in blue and cytoplasm in red. Panels B and D show the corresponding virtual trichrome images generated from DUET, replicating the color contrast, structural detail, and visualization of the real trichrome. Panels A and B are shown at low magnification, and panels C and D at high magnification.(A–B) Scale bar: 200 μm. (C–D) Scale bar: 50 μm.

### Hybrid Virtual Stain Overlay

A novel application of virtual stains generated from DUET is the creation of hybrid virtual stains. Figure 6 shows original brightfield H&E images at low (A) and high (C) magnification alongside hybrid virtual stains of the same fields (B, D). The hybrid stain consists of an overlay of the colorized DUET-extracted collagen signal onto the H&E image in a pixel-registered fashion. This view can be toggled in the software, enabling pathologists to reference collagen distribution directly within the context of the original H&E architecture. This approach enables better visualization and distribution of collagen fibers, while leveraging the strengths of the H&E stain (e.g., nuclear and cytoplasmic detail).

**Figure 6.**
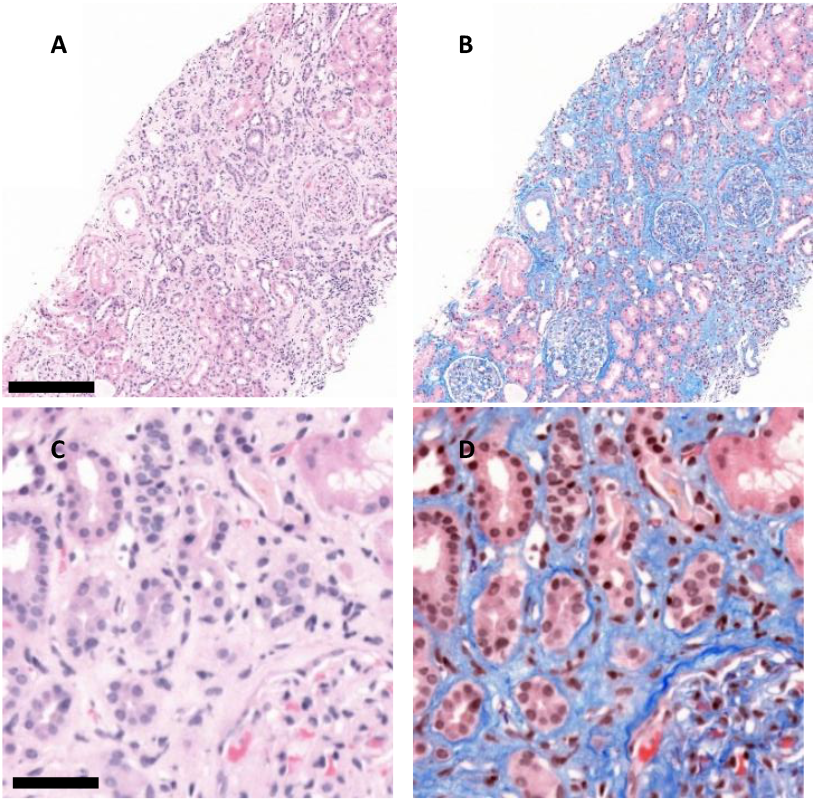
Low- and high-power comparisons of original H&E and hybrid H&E images from the same kidney biopsy region. Panels A and C display the conventional brightfield H&E images at low and high magnification, respectively. Panels B and D show the corresponding hybrid H&E images with a pixel-registered DUET collagen overlay, enabling direct visualization of collagen distribution within the native H&E architecture. (A–B) Scale bar: 200 μm. (C–D) Scale bar: 50 μm.

### Pathologist Interstitial Fibrosis Estimation of Real and Virtual Trichrome Stains

To prove the utility of DUET-derived virtual trichrome stains, four renal pathologists visually estimated the extent of IF in a set of 32 real trichrome whole slide images and their corresponding virtual trichrome whole slide images. The detailed estimates are included in Figure 7. While the absolute IF estimates tend to be slightly higher with real trichrome WSIs, the overall trends for each pathologist remain consistent across both modalities.

**Figure 7.**
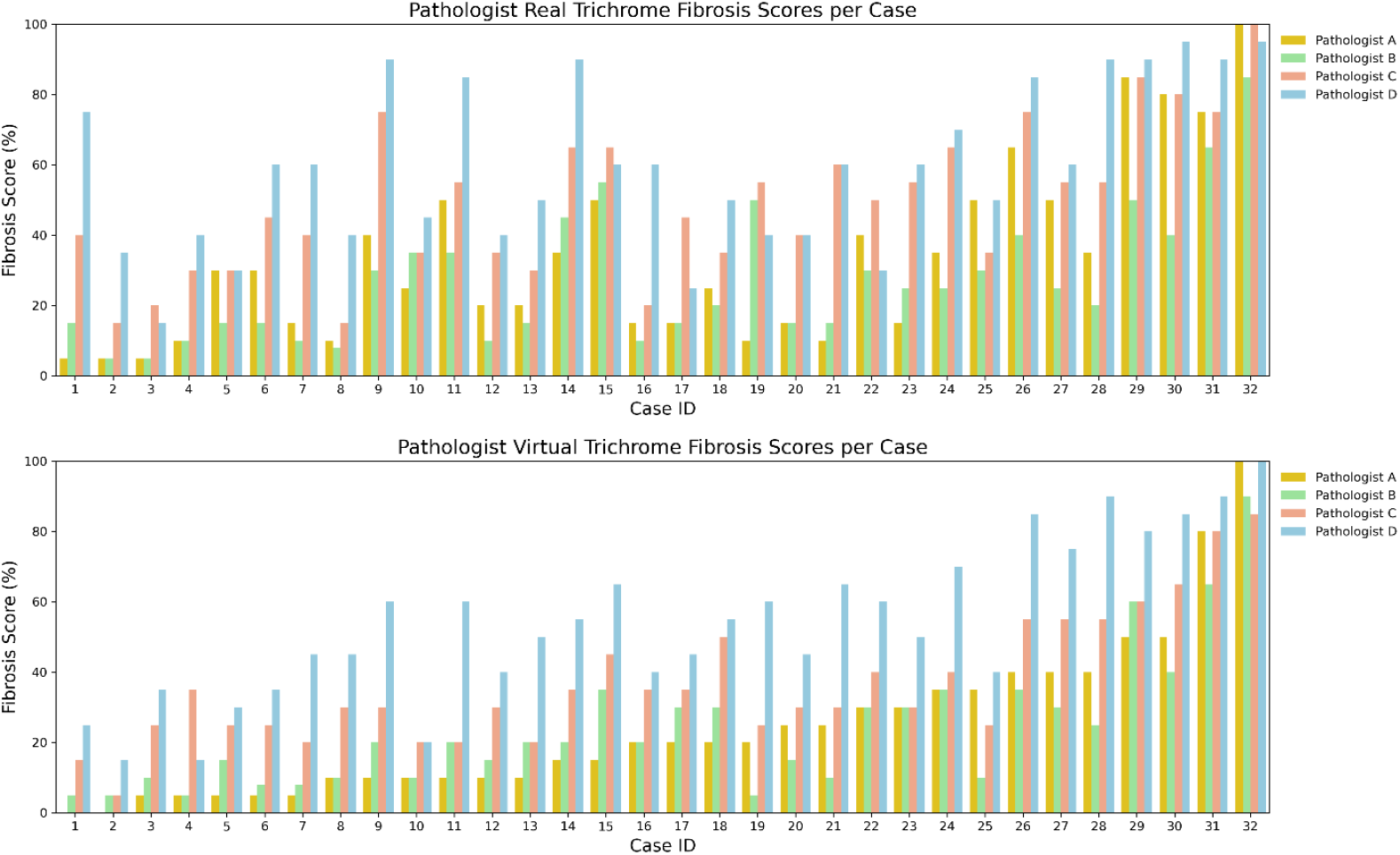
Bar plots of fibrosis scores assigned by four pathologists for each case, comparing real trichrome (top) and virtual trichrome (bottom) images. Each bar represents a single case, with individual pathologist scores shown in a different color. Cases are ordered along the x-axis from left to right based on ascending fibrosis scores assigned by Pathologist A in the virtual trichrome condition.

Intraclass correlation coefficients (ICCs) were calculated to assess the agreement of fibrosis estimation for each pathologist when using the real and virtual trichrome WSIs. As seen in Table 2, intra-pathologist ICCs demonstrate moderate to substantial agreement, indicating that the two modalities are comparable for IF estimation. ICCs were also calculated across all four pathologists based on using either real or virtual trichrome WSIs (Table 2). A higher inter-pathologist agreement was observed when using virtual trichrome (ICC = 0.536) compared to real trichrome (ICC = 0.477) WSIs, indicating that IF estimation consistency between pathologists is improved when using DUET-generated virtual trichrome WSIs.

**Table 2.** Intraclass correlation coefficient (ICC) by subgroup (pathologist and stain), with 95% confidence interval bars.

| Category | ICC (95% CI) |
| --- | --- |
| Pathologist |  |
| A | 0.673 (0.425, 0.827) |
| B | 0.767 (0.570, 0.878) |
| C | 0.740 (0.548, 0.864) |
| D | 0.562 (0.265, 0.759) |
| Stain |  |
| Trichrome | 0.477 (0.267, 0.636) |
| vTrichrome | 0.536 (0.338, 0.684) |

Similarly, when each pathologist’s individual IF estimates were compared to the average estimate across all four pathologists (used here as a reference standard), the coefficients of determination (R^2^) when using virtual trichrome WSIs were consistently higher than those when using real trichrome WSIs, further supporting improved alignment with the consensus when using the virtual trichrome WSIs (Table 3).

**Table 3.** Coefficients of determination (R^2^) between individual pathologists’ fibrosis scores and the average score across all pathologists, for both real and virtual trichrome images. Higher R^2^ values in the virtual trichrome condition indicate greater agreement with the consensus.

| Pathologist | Real Trichrome ($R^2$) | Virtual Trichrome ($R^2$) |
| --- | --- | --- |
| A | 0.73 | 0.84 |
| B | 0.78 | 0.88 |
| C | 0.85 | 0.92 |
| D | 0.88 | 0.91 |

### Computational Collagen Quantitation of DUET Images

Collagen images extracted with DUET were used to computationally quantify the degree of IF in WSIs, which was defined as the proportion of collagen-positive pixels within the interstitium relative to the total cortical area. Of note, this definition does not equate to the pathologists’ estimation of IF given that pathologists include tubules within the areas they consider involved by IF. Glomeruli, large caliber arteries, and non-cortical areas were manually excluded for the computational IF quantitation. These computational quantifications were compared to the average IF estimates of the renal pathologists (Figure 8). The coefficient of determination (R^2^) between pathologists and computational IF quantitation was higher for virtual trichrome WSIs (R^2^ = 0.44) than for real trichrome WSIs (R^2^ = 0.29), indicating that our computational approach shows a greater alignment with the virtual trichrome modality.

**Figure 8.**
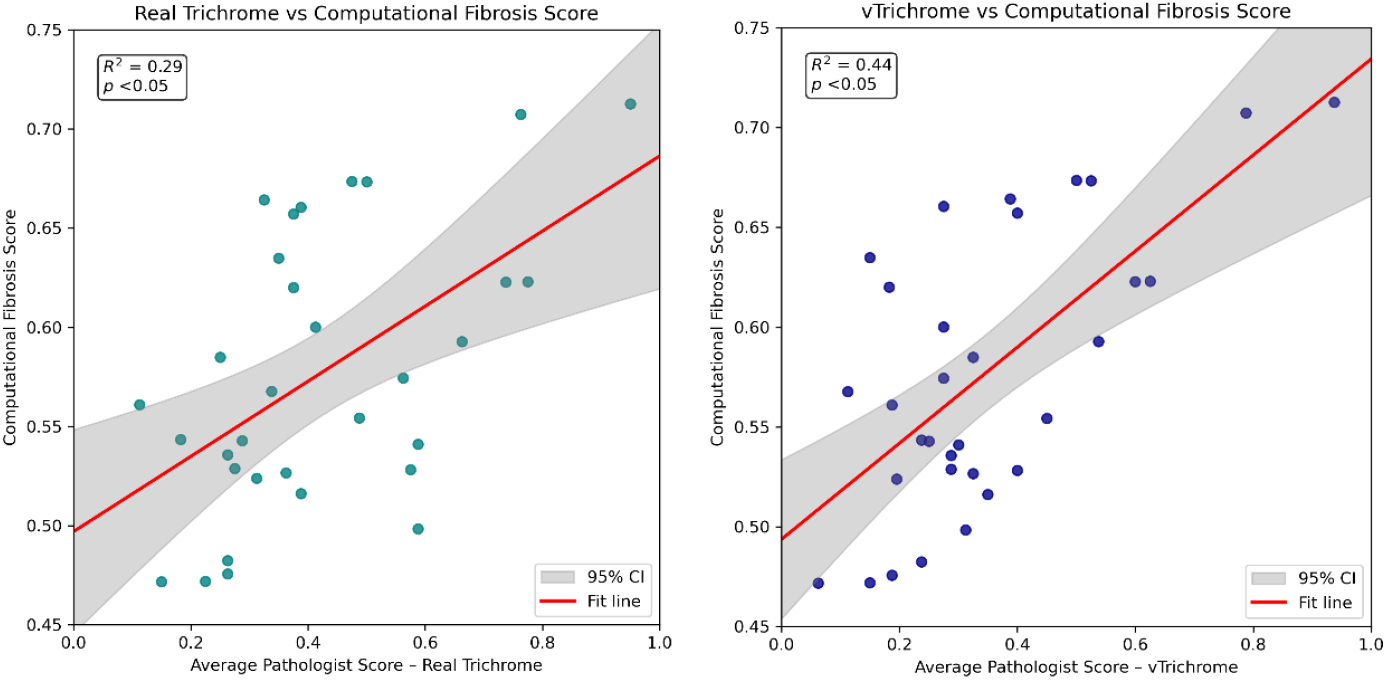
Scatter plots comparing the computational fibrosis score to the average manual pathologist score for each case, shown for real trichrome (left) and virtual trichrome (right). Each point represents a single case; the red line represents the least-squares fit, and the gray band indicates the 95% confidence interval. Correlation with the consensus was higher for vTrichrome (R^2^ = 0.44, p < 0.05) than for real trichrome (R^2^ = 0.29, p < 0.05).

## DISCUSSION AND CONCLUSION

This study demonstrates the potential of DUET imaging to generate clinically useful virtual trichrome stains by extracting collagen-specific fluorescence signals from H&E-stained slides, providing a quantitative framework for automated evaluation of kidney fibrosis and laying the groundwork for future patient outcome prediction. This is achieved through a novel imaging approach that offers an accurate and reproducible means of assessing interstitial fibrosis in renal biopsies, closely mimicking traditional trichrome stains while improving scoring agreement and enabling computational quantification.

Virtual trichrome stains visually recapitulate key histopathological features observed in traditional Masson’s trichrome stains, including collagen deposition patterns associated with IF. The virtual trichrome WSIs demonstrate high reproducibility in qualitative stain consistency, even when input H&E stains exhibit variable quality, likely contributing to the higher intra-pathologist agreement observed in this study. Importantly, virtual trichrome WSIs are generated from the same section as the H&E slide rather than an adjacent section, eliminating the risk of missing focal lesions that may not be present in serial sections.

DUET also enables the creation of hybrid WSIs featuring a toggleable collagen overlay on the original H&E stain. These hybrid views leverage the nuclear and cytoplasmic detail of H&E with the collagen fiber visualization of traditional trichrome stains. Once integrated into a digital pathology workflow, hybrid stains have the potential to enhance both efficiency in routine pathology workflows and diagnostic confidence, similar to tissue windows in radiology, which are applied to computed tomography scans.

Given that DUET isolates the collagen signal under fluorescence illumination, we used this signal to generate computational estimates of IF degree. These quantitative estimates showed moderate correlation with pathologists’ visual estimates, suggesting viability of this computational approach. The computational fibrosis score correlated more strongly with virtual trichrome estimates than with real trichrome estimates, likely reflecting their shared source material and differences in tissue sectioning between H&E and serial trichrome slides. The moderate correlation with pathologist estimates in both modalities highlights the computational approach’s potential to align with expert interpretation. Although we manually excluded non-tubulointerstitial structures for this study, further integration of automated segmentation tools would enable fully-automated IF evaluation.

A limitation of our study is the lack of a widely accepted “gold standard” for IF. Although the pathologists’ visual estimation is often used, this type of subjective evaluation is plagued by both inter- and intra-pathologist variability. The four pathologists in this study are from three different academic institutions and trained at four different training programs, which likely contributed to some degree of intra-pathologist variability. Thus, we used the average of the pathologists’ estimates as a “consensus” for comparison in most of the analyses. We also used second harmonic generation, an accepted method for collagen imaging, to establish a “gold standard” for the collagen signal extracted from DUET.

Taken together, these results suggest that DUET imaging, particularly when paired with computational analysis, holds promise as a reproducible and scalable adjunct to traditional histology. Future prospective studies should explore its impact on clinical outcomes, as well as its integration with digital workflows for remote, frozen section, or high-throughput pathology settings.

## Data availability

Data underlying the results presented in this paper are not publicly available at this time but may be obtained from the authors upon reasonable request.

